# A Wide-bandwidth Nanocomposite-Sensor Integrated Smart Mask for Tracking Multi-phase Respiratory Activities for COVID-19 Endemic

**DOI:** 10.1101/2022.03.28.22273021

**Authors:** Jiao Suo, Yifan Liu, Cong Wu, Meng Chen, Qingyun Huang, Yiming Liu, Kuanming Yao, Yangbin Chen, Qiqi Pan, Xiaoyu Chang, Ho-yin Chan, Guanglie Zhang, Zhengbao Yang, Walid Daoud, Xinyue Li, Roy Vellaisamy, Xinge Yu, Jianping Wang, Wen Jung Li

## Abstract

A global sentiment in early 2022 is that the COVID-19 virus could become endemic just like common cold flu viruses soon. The most optimistic view is that, with minimal precautions, such as vaccination, boosters and optional masking, life for most people will proceed as normal soon. However, as warned by A. Katzourakis of Oxford University recently [1], we must set aside lazy optimism, and must be realistic about the likely levels of death, disability and sickness that will be brought on by a ‘COVID-19’ endemic. Moreover, the world must also consider that continual circulation of the virus could give rise to new variants such as the new BA.2 variant (a subvariant of Omicron) continues to spread across the US and parts of Europe. Data from the CDC is already showing that BA.2 has been tripling in prevalence every two weeks [2]. Hence, globally, we must use available and proven weapons to continue to fight the COVID-19 viruses, i.e., effective vaccines, antiviral medications, diagnostic tests and stop an airborne virus transmission through social distancing, and ***mask wearing***. For this work, we have demonstrated a smart mask with an optimally-coupled ultra-thin flexible soundwave sensors for tracking, classifying, and recognizing different respiratory activities, including *breathing, speaking*, and two-/tri-phase *coughing*; the mask’s functionality can also be augmented in the future to monitor other human physiological signals. Although researchers have integrated sensors into masks to detect respiratory activities in the past, they only based on measuring temperature and air flow during coughing, i.e., counting only the number of coughs. However, coughing is a process consisting of several phases, including an explosion of the air with glottal opening producing some noise-like waveform, a decrease of airflow to decrease sound amplitude, and a voiced stage which is the interruption of the air flow due to the closure of glottal and periodical vibration of partly glottis, which is not always present. Therefore, sensors used for cough detection should not be only sensitive to subtle air pressure but also the high-frequency vibrations, i.e., a pressure sensor that needs to be responsive to a wide input amplitude and bandwidth range, in order to detect air flows between hundreds of hertz from breath, and acoustic signals from voice that could reach ∼ 8000 Hz. Respiratory activities data from thirty-one (31) human subjects were collected. Machine learning methods such as Support Vector Machines and Convolutional Neural Networks were used to classify the collected sensor data from the smart mask, which show an overall macro-recall of about 93.88% for the three respiratory sounds among all 31 subjects. For individual subjects, the 31 human subjects have the average macro-recall of 95.23% (ranging from 90% to 100%) for these 3 respiratory activities. Our work bridges the technological gap between ultra-lightweight but high-frequency response sensor material fabrication, signal transduction and conditioning, and applying machining learning algorithms to demonstrate a reliable wearable device for potential applications in continual healthy monitoring of subjects with cough symptoms during the eventual COVID-19 endemic. The monitoring and analysis of cough sound should be highly beneficial for human health management. These health monitoring data could then be shared with doctors via cloud storage and transmission technique to help disease diagnosis more effectively. Also, communication barriers caused by wearing masks can be alleviated by combining with the speech recognition techniques. In general, this research helps to advance the wearable device technology for tracking respiratory activities, similar to an Apple Watch or a Fitbit smartwatch in tracking physical and physiological activities.

## Introduction

Since the emergence and pandemic of the coronavirus disease in 2019 (COVID-19) [3], it has been generally accepted that universal masking is a necessary measure against the worldwide spread of COVID-19 since wearing masks can effectively prevent the transmission of coronavirus and influenza viruses from infected individuals [4-6]. Many countries lay down the laws requiring the use of masks [7, 8], and wearing mask has become a daily necessity, even being part of people’s social life. In fact, as recent as February 2022, the Centers for Disease Control and Prevention (CDC), USA, has published a study on the effectiveness of face mask or respirator use in indoor public settings for prevention of SARS-CoV-2 infection, which indicates consistent use of a face mask or respirator in indoor public settings was associated with lower odds of a positive SARS-CoV-2 test result, i.e., the study found that there was a 66% lower odd for people wearing surgical masks and 83% lower odds for people wearing N95/KN95 respirator to test positive for COVID-19 than people who did not [9].

Across the globe, there has been a sentiment in early 2022 that the Covid-19 virus could become just endemic just like common cold flu viruses soon. The most optimistic view is that, with minimal precautions, such as vaccination, boosters and optional masking, life for most people will proceed as normal soon. For example, some places are already planning for the endemic stage. Spanish Prime Minister Pedro Sanchez has asked European officials to discuss when COVID-19 should be considered endemic [10], and California has adopted the USA’s first endemic policy for COVID [11], which emphasizes on the prevention and quick reaction to outbreaks instead mandated masking. However, as warned by A. Katzourakis of Oxford University recently[1], we must set aside lazy optimism, and must be realistic about the likely levels of death, disability and sickness that will be brought on by a ‘COVID-19’ endemic. It is important to remember that endemic does not correspond to harmlessness. Malaria, for example, has become widely recognized in ancient Greece by the 4th century BC, but is still considered endemic in 87 countries worldwide as reported by CDC in 2021 [12]. In fact, still nearly half the world’s population lives in areas at risk of malaria transmission and malaria caused an estimated 241 million clinical cases and 627,000 deaths in 2020. Moreover, the world must also consider that circulating virus could give rise to new variants such as the new BA.2 variant (a subvariant of Omicron) continues to spread across the US and parts of Europe. Data from the CDC is already showing that BA.2 has been tripling in prevalence every two weeks [2]. Hence, globally, we must use available and proven weapons to continue to fight the COVID-19 viruses, i.e., effective vaccines, antiviral medications, diagnostic tests and stop an airborne virus transmission through social distancing, and ***mask wearing***.

Therefore, deploying a face mask to monitor human physiological signals has been highly beneficial for personal and public health [13-27]. Different kinds of smart masks have been proposed to detect body signals including respiratory rate/heart rate [13-20], skin temperature [14, 18, 21-25], cough counting (based on temperature and pressure change) [18, 20], blood oxygen [14] and airborne pathogens [26, 27]. Among them, respiratory activities, in particular cough as a key symptom of respiratory illness, are usually of great significance in assisting the diagnosis of diseases such as pertussis and asthma [28-30], and the latest research also further indicates that the impact of COVID-19 on the respiratory system may lead to notable changes in the voice of infected people which can be determined by speak, breath, and cough [31-33]. Therefore, it is highly desirable to develop smart masks to monitor respiratory activities such as *breath, cough*, and *speech*.

As one of the most important respiratory activities, breath-related signals (e.g., heart rate and breath rate) are usually detected by off-the-shelf photoplethysmography (PPG) sensor [14, 15] and thermistor [18] integrated with masks. In addition to the commercial ones, advanced nanogenerators and flexible sensors based on new nanomaterials are also used recently. It is known that breath is the process of air flowing in and out of the lungs namely exhalation and inhalation [34]. Therefore, it also can be detected as air-flow-driven pressure. For example, the nanogenerators based on the nanofibrous [13] and the nanostructured polytetrafluoroethylene (n-PTFE) thin film [16] were used to detect the breath activities successfully. An ultrathin pressure sensor with piezoelectric-like properties was also integrated in a face mask to detect human breath activities [20]. As for coughing, researchers used integrated sensors detect the temperature change [18] and air flow [20] during coughing. These coughing detection sensors focus only on cough counting, the obtained information would be limited since coughing is a process consisting of several stages. Coughing initiated as a series of respiratory activities to cause a sudden explosion of air along with the cough sound which usually consists of three phases [35-38]. The first stage is an explosive of the air with glottal opening producing some noise-like waveform, and then comes to the second steady stage when the airflow is decreased causing the sound amplitude is also decreasing. The third is the voiced stage which is the interruption of the air flow due to the closure of glottal and periodical vibration of partly glottis, which is not always presented [38]. Therefore, sensors used for cough detection should not be only sensitive to subtle air pressure but also the high frequency vibrations.

Another function of the smart mask that draws attention is the speech detection. The speech is produced by the vocal folds vibration induced air vibration when vocal folds come close as the air passing through during the exhalation of air from the lung [39]. Although a few research also suggest medical masks have no negative effect on speech understanding [40], more recent investigations have further demonstrated that both standard surgical masks and N95/KN95 respirators have an influence on the acoustic characteristics of voice [41-44], attenuating the mean spectral level by 2.0–5.2 dB in the high-frequency region from 1 to 8 kHz while not significantly affecting the low-frequency range from 0 to 1 kHz [41]. Additionally, the masks with higher barrier level have been proved to have greater attenuation of high-frequency components during the transmission of sound [41, 42], and masks impact more on human speech recognition in a higher level of background noise [45]. The high-frequency signals, generated by vocal cord vibration [46], provide perceptual information for individual speaker gender [47, 48] and contribute to speech intelligibility [49], thus, wearing masks causes the dampening of high-frequency spectral energy which leads to decline in clarity of speech, bringing trouble to daily life. For example, mobile phone software that requires voice recognition cannot work properly in noisy environments when a person is wearing a mask, and the health monitoring related to the human voice will also be affected. Therefore, integrating a ‘voice recorder’ inside of the mask would be beneficial for speech recognition and understanding when wearing a face mask. What’s more, human speech also provides health information and can help disease diagnose [50-52].

Based on the above facts, for monitoring the three common respiratory activities (i.e., breath, cough, and speech), this work presented a novel smart mask based on an ultra-thin sponge structure-based sensor made of CNT/PDMS nanocomposites with an easy fabrication process but high sensitivity in both static pressure and dynamic pressure ranges. The 400μm thick sponge structure shows a static pressure sensitivity of about 0.85kPa^-1^ and responds to high-frequency dynamic pressure generated by human voice, i.e., sound harmonic energy up to 4000Hz. Air pressure caused by air movements consisting of air directional flow and air vibration can also be detected, and their typical features were investigated. We report here a smart mask by integrating flexible high-frequency response sensors on commercial masks to monitor human respiratory sounds (including breath, cough, and speech). Different characteristics of the three different respiratory activities were successfully captured. Thirty-one human subjects were recruited to collect the respiratory activity data of breath, cough, and speech. These data were further processed and classified by Support Vector Machines (SVM)/Convolutional Neural Networks (CNN), which has an over macro-recall of about 91.43%/93.88% for the three respiratory sounds among all 31 subjects. For individual subjects, all the 31 human subjects have the macro recall above 90% (with the maximum as high as 100%) and the average reaches 95.23% for these 3 different types of sounds. Several applications could be developed using this smart mask. For example, as shown in Figure 1, the important health related signals, respiratory rate (and further heart rate which is about 4 times of respiratory rate during rest) and cough can be recorded daily. As a typical respiratory disease symptom, cough is one of the commonest reasons for consultation with a doctor [53]. The monitoring and analysis of cough sound should be highly beneficial for human health management. These health monitoring data could then be shared with doctors via cloud storage and transmission technique to help disease diagnosis more effectively. Also, communication barriers caused by wearing masks can be alleviated by combining with the speech recognition techniques. In general, this research helps to advance the wearable device technology for tracking respiratory activities, similar to an Apple Watch or a Fitbit smartwatch in tracking physical and physiological activities. The smart mask discussed here could eventually incorporate other physiological tracking capabilities, and wireless telecommunication for cloud-based data analytics to achieve respiratory disease early warning and tracking. For example, with continual development, this smart mask can be used to monitor patients with Chronic Obstructive Pulmonary Disease (COPD) which is a persistent inflammatory lung disease that causes obstructed airflow from lungs and could include patients with post-COVID-19 symptoms. COPD could severely affect patients’ quality of life and may also cause significant social and economic burdens, and as a progressive and irreversible disease, the major symptoms of COPD include breathing difficulty, cough, mucus (sputum) production and wheezing. However, many COPD patients are under-diagnosed, which lead to the delay of treatment until later stage or during acute exacerbation period. Therefore, the tracking, classification, and treatment of coughs are extremely important. On the other hand, recent research results indicate that COVID-19 patients with respiratory infections may lead to unique changes in the sound when they *speak, breathe*, and *cough*. These subtle acoustic differences are indistinguishable to the human ear, but are discernible by AI algorithms with an accuracy of more than 95% (e.g., see [33]). In general, the recognition of sounds from speaking, breathing, and coughing has tremendous potential for detecting not just COVID-19 but for other COPD symptoms as well. Currently, many researchers are collecting cough audio samples from the public to build AI models and improve machine learning algorithms through smartphone apps and web-based platforms. However, from a hygiene point of view, coughing or speaking on open surfaces is not desirable, as it may result in further transmission of the respiratory diseases. For this work, we have demonstrated a smart mask with an optimally-coupled ultra-thin flexible soundwave sensors for tracking, classifying, and recognizing different respiratory activities, including *breathing, speaking*, and *coughing*; the mask’s functionality can also be augmented in the future to monitor other human physiological signals. Ultimately, we envision the smart mask as a wearable device that can continuously track a patient’s daily respiratory activities, and which can be used to facilitate the screening and diagnosis of cough-related diseases.

**Figure 1.**
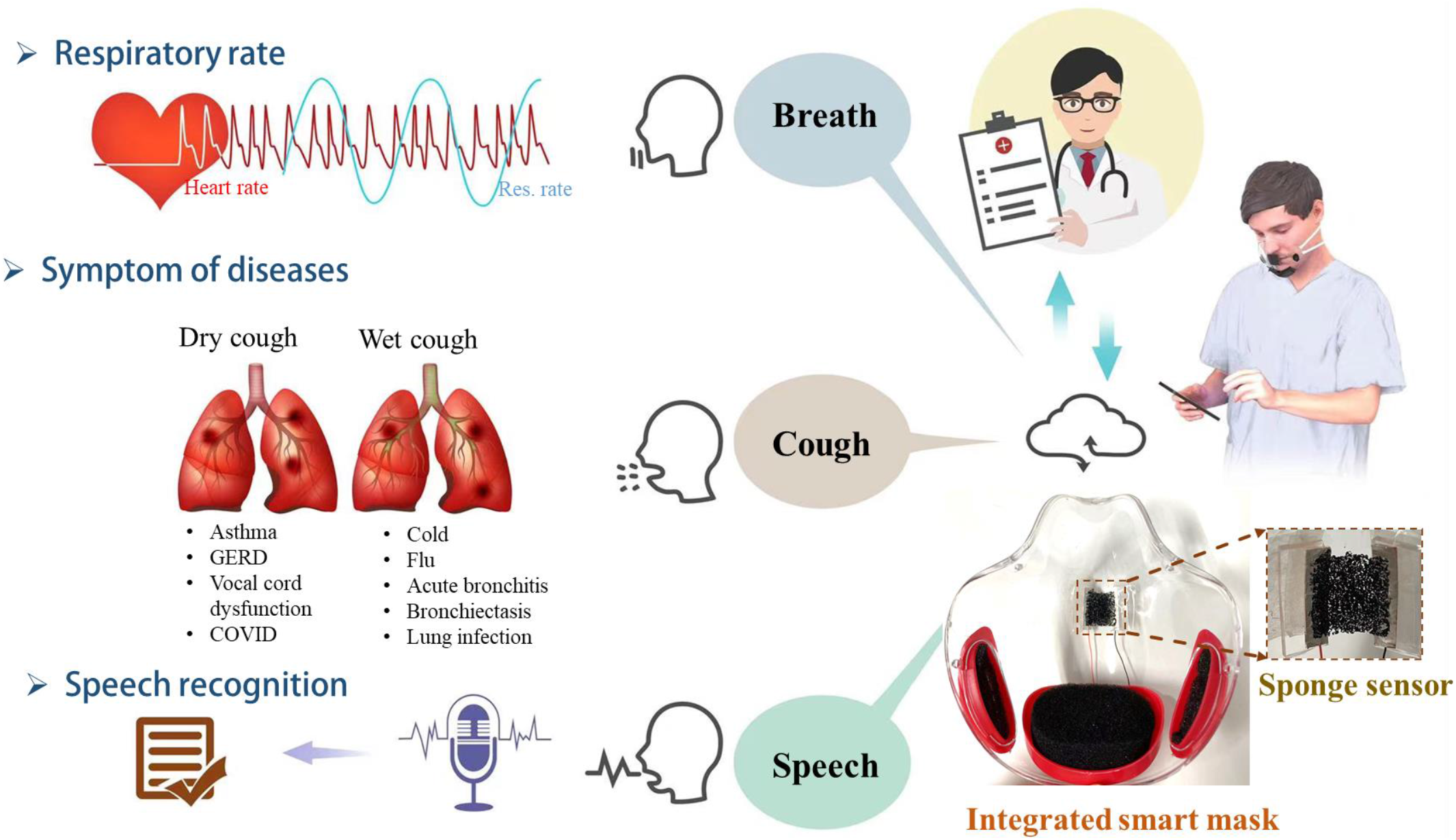
Application of smart mask monitoring human daily respiratory activities

## Methods

### Preparation of the CNT/PDMS nanocomposites

The preparation process for the CNT/PDMS composites solution was by mixing multi-wall CNTs (MWCNTs; XFNANO, China) and PDMS (Sylgard184; Dow, USA) in isopropyl alcohol (IPA; Anaqua Chemical Supply, Hong Kong). IPA was used as the solvent because both CNT and PDMS are partially soluble in it [54, 55]. The MWCNTs have diameters of 10–20 nm and lengths of 10–30 μm (provided by the manufacturer). First, MWCNTs were dispersed in a sufficient quantity of IPA and ultrasonicated for 20 min to obtain a dispersion of CNTs. Then, PDMS-A base was added into the dispersion and ultrasonication for 10 min. Subsequently, the mixture was placed on a hotplate (IKA, Germany) maintained at 55°C to completely evaporate the IPA. Thereafter, PDMS-B agent (the weight ratio of PDMS-A and PDMS-B is 10:1) was added to the solution and mechanically mixed. Finally, air bubbles were removed from the mixture through vacuum treatment. In this manner, CNT/PDMS composites with CNT concentrations of 2wt%, 2.5wt%, 3wt%, 4wt%, and 5wt% were prepared by mixing 0.2/0.25/0.3/0.4/0.5g of MWCNTs with 10g PDMS.

### Preparation of the CNT/PDMS sponge

CNT/PDMS sponges were fabricated using sacrificial sugar cubes (Taikoo; purchased from a supermarket in Hong Kong) measuring 19.6 mm × 18.4 mm ×11.7 mm (thickness). The CNT/PDMS composite solution was dropped (with syringe) on to the surface of a sugar cube and spread smoothly across its surface. Then, a second sugar cube was placed atop the first cube and cured in an oven at 70°C for about 2h. Finally, the remaining sugar was dissolved in deionized water to obtain a CNT/PDMS sponge sheet. This modified imprinting technique was used to fabricate sponges with a very thin thickness of about 400 μm. Sponge samples with different CNT weight concentrations (2wt%–5wt%) were prepared from the same batch of sugar cubes, and all samples were made the same thickness by controlling the solution amount.

### Fabrication of CNT/PDMS sponge-based pressure sensor

Then, the flexible pressure sensors were fabricated based on the thin film sponge. Two sheets of copper tape served as the electrodes and were bonded to electrical wires. The sponge was attached to the electrodes using conductive silver paste, after which the paste was solidified using a hot air gun (Saike, China) at 100°C for approximately 1 min. A piece of parafilm was served as a substrate when needed. The entire sensor could also be packaged by coating a layer of parylene-c thin film on the surface. The coating process was applied under 17 miliTorr, and the pyrolysis temperature was 690°C, vaporization temperature was 175°C, deposition temperature was at room temperature. The resulting thickness was about 30nm when 0.1g parylene-c were applied.

### Characterization of CNT/PDMS sponge-based sensor

The porous structure of the CNT/PDMS sponge was characterized using scanning electron microscopy (SEM, FEI Quanta 450). Direct current resistance was measured using a digital multimeter (Fluke 15B+, USA), and impedance characteristics were recorded using an impedance analyzer (HIOKI IM 3570, Japan) in the 4 Hz– 5 MHz range at 1 V. The equivalent circuit models of the CNT/PDMS sponge were extracted using an EIS spectrum analyzer (open-source software). A series of static pressures from 27.9 Pa to 2.5 kPa were realized by applying different weights. The changes in impedance under different frequencies (4 Hz, 10 kHz, 1 MHz, 2.5 MHz, and 5 MHz) were recorded using the impedance analyzer at a sampling rate of 1 Hz. Also, the vibration input with 100∼800Hz was provided by a vibration speaker, while the vibration acceleration was measured by the laser doppler vibrometer (LDV). The measurement voltage divider circuit contained a DC power source (5 V) and a voltage divider resistor. The change in the output voltage signal of the sensor was recorded using an oscilloscope. Then, the corresponding resistance change was calculated. At last, the sensor was fixed freestanding. Air directional flow and air vibration were applied from a distance. The air directional flow pressure was provided by an air gun blowing air to the sensor with constant velocity, and the air vibration was induced by the speaker playing a 315Hz sound in front of the sensor. The measurement circuit is based on the the microphone pre-amplifier board MAX4466 with replacing the integrated electret with our self-developed sensor.

### Smart mask preparation and human respiratory activities detection

The developed thin film sponge-based sensor was then fixed to the inner side of a commercial transparent mask. The commercial polycarbonate mask used in this project has a hard texture (with a Young’s modulus of about 2.4GPa for the polycarbonate [56]) and can keep a fixed shape with three filters located on the two sides of the cheeks, and the chin, respectively, as shown in Fig. 1. The sensor was fixed to the approximate position of the mouth and nose of the mask. Firstly, the two pieces of sensor electrodes was attached to two pieces of PDMS which has a similar area of the electrodes and about 2mm thick with double-sided tape. Then, the sensor was fixed to the inside of the mask by attaching the two pieces of PDMS to the mask with double-sided tape to make the sponge sensing area flat and freestanding. A wireless data acquisition board was also dedicated based on the ESP32, which is a feature rich MCU with integrated Wi-Fi and Bluetooth connectivity. It collects data with 8kHz sampling rate and transmits data to a computer via WIFI. This homemade ESP32-based wireless module has the size of 31 mm x 22 mm x 14 mm and weight of 13g (with battery). The power consumption of this device is low, in which the current is 56mA at idle status (i.e., the device is standby and ready for data collection) and 140mA at working status. Volunteers were asked to wear the mask naturally, and perform the respiratory activities of breath, cough (voluntary) and speech (i.e., speaking ‘robot’). Six pieces of data were obtained for each human each activity, while each piece of data last for 40 seconds and consist of at least five breath/cough/speech signals. The human subjects consist of male (10) and female (20), and the age is between 24 and 32.

### Data process and classification

Signals of human respiratory activities were converted to audios by a self-developed program, followed by a noise reduction work. A segmentation process was performed manually to select the clean signals for each activity from the noise according to the visualized waveform and spectrogram. Fifty-three features (shown in **Table1**) were extracted for the classification process. Before the final classification, the principal component analysis (PCA) was used to reduce the dimension and compute the main components of all the features. Support vector machine (SVM) algorithm was applied to perform the classification (50% data for training while 50% for testing) and the metric recall was selected to evaluate its performance. Recall predicts the correct proportion of all samples that are actually positive, the recall for activity *i* is defined as:

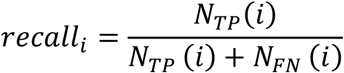

**Table 1.** The 53 features extracted from each segmented signal

| 53 features |  |
| --- | --- |
| Feature | Number |
| Signal length | 1 |
| Mean fundamental frequency | 1 |
| Harmonic-noise-ratio (HNR) | 1 |
| FFT Max Amplitude | 1 |
| Frequency of FFT Max Amplitude | 1 |
| Kernel Distribution Estimation (KDE) coefficient | 1 |
| Power spectral density (PSD) variance | 1 |
| Average spectrum energy | 1 |
| Spectrum energy variance | 1 |
| Sub-band Spectrum energy (0~250Hz, 250~500Hz, 500~1k Hz, 1k~2k Hz and 2k~4k) | 5 |
| Mel Frequency Cepstral Coefficients (MFCCs) | 39 |

A high recall value indicates that most of the behavior samples are correctly classified, due to the size differences between three activities, we adapt the average recall value (macro-recall) as the main metric to represent the model overall performance, which is expressed as:

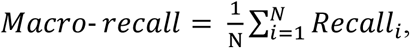

where *N* is the total number of activities.

## Results

### Large-area, ultra-thin, porous sponge structure

Figure 2**a** illustrates the preparation process of the nanocomposites of CNT/PDMS by mixing PDMS with MWCNT while IPA applied as the solvent. This mixture solution can be processed into different structures with templates and curing process. Figure 2**b** presents the fabrication of the thin film sponge structure with the CNT/PDMS materials using a novel modified imprint technique. The porous sponge has a minimum thickness of about 400μm (area size of 19.6 mm ×18.4 mm, the same as the sugar template) with high flexibility (Figure 2**c**). It has a varied pore size in the range of about 150μm∼550μm. Two pieces of soft copper tape served as the electrodes are bonded with the thin sponge structure with silver paste, so that this sensor can be connected to a circuit to study its electrical properties and sensing performance.

**Figure 2.**
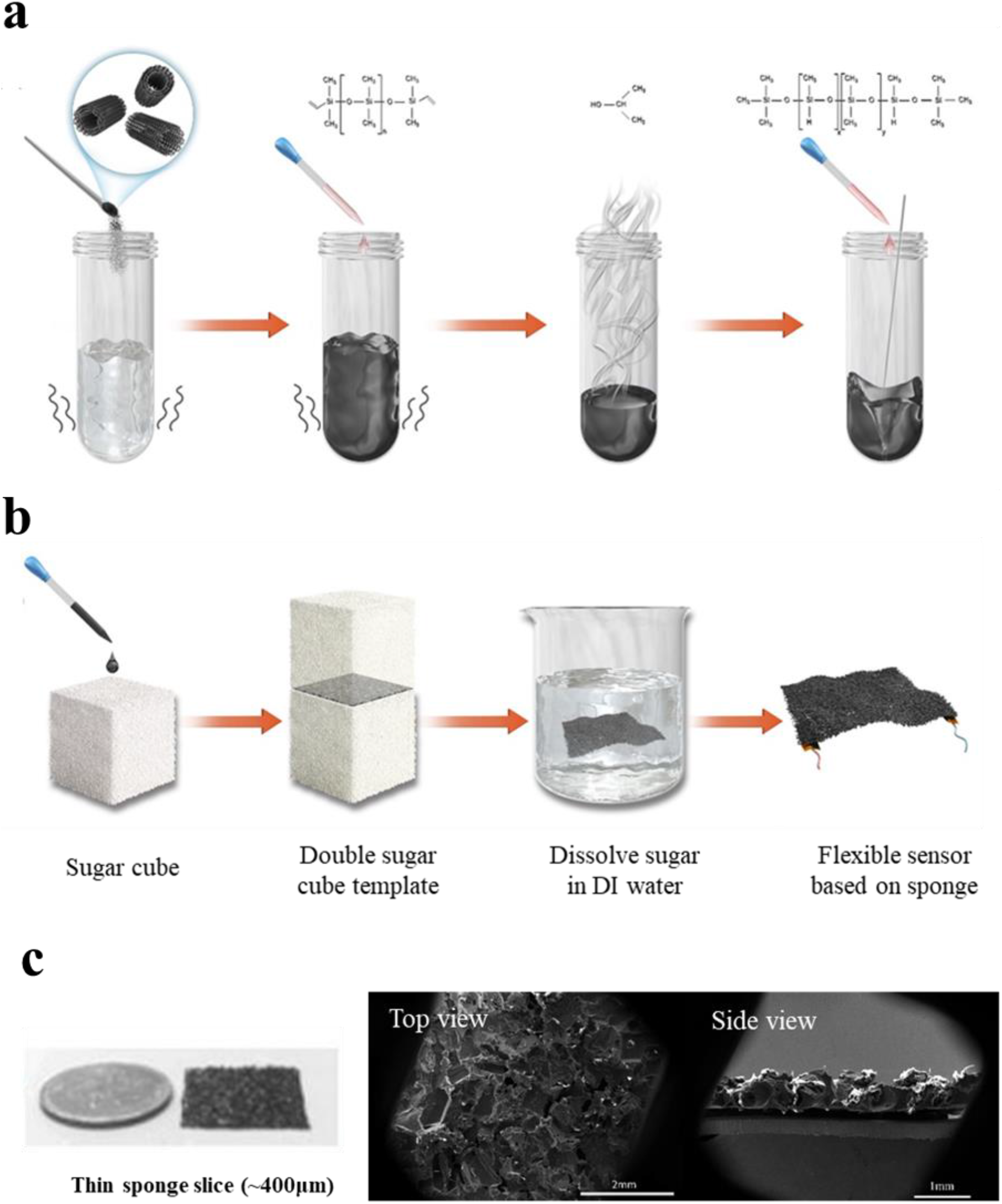
Schematic showing the flexible sensor fabrication. (a) preparation of CNT/PDMS nanocomposites, and (b) CNT/PDMS sponge and sponge-based sensor. (c) Illustration of the sponge-based sensor and SEM images to show the porous structure.

### Electrical properties of the CNT/PDMS sponge-based sensors

This kind of synthetic nanocomposites by mixing polymer and nano conductive materials usually have complex electrical properties depending on the ratio of the conductive materials and cannot be treated as a capacitor or a resistor simply [57-60]. Therefore, it is necessary to study the electrical properties of the prepared CNT/PDMS sponge (∼400μm) sensors with different CNT concentrations in detail to lay the foundation for its application. Figure 3**a** shows the direct current resistance values (R_DC_) of the sensor with different CNT content. A higher CNT concentration is associated with a lower resistance of the CNT/PDMS sponge. The resistance can reach hundreds of thousand ohms when the CNT content is 2wt%. In addition, the R_DC_ exhibited slight change at CNT concentrations of 3wt% or higher. Regarding the impedance properties, the changes in the phase angle (θ) and total impedance (Z) of the CNT/PDMS sponge-based sensors at varying frequencies from 4 Hz–5 MHz are shown in Figure 3**b, c**. At relatively low frequencies, resistive behavior was dominant. The total impedance did not change with frequency, and the phase angle remains near zero. As the frequency increased, both the total impedance and phase angle changed. At high frequencies, the phase angle was no longer zero, indicating that the sponge had both the resistance component (i.e., the real component of Z) and the reactance component (i.e., the imaginary component of Z). Accordingly, the test frequency range could be delineated into resistive and capacitive regions, which respectively increased and decreased with increasing CNT concentration. In addition, the sponges with CNT concentrations of 3wt%, 4wt%, and 5wt% exhibited similar impedance curves (frequency *vs* θ and Z) and they behave more like a resistor.

**Figure 3.**
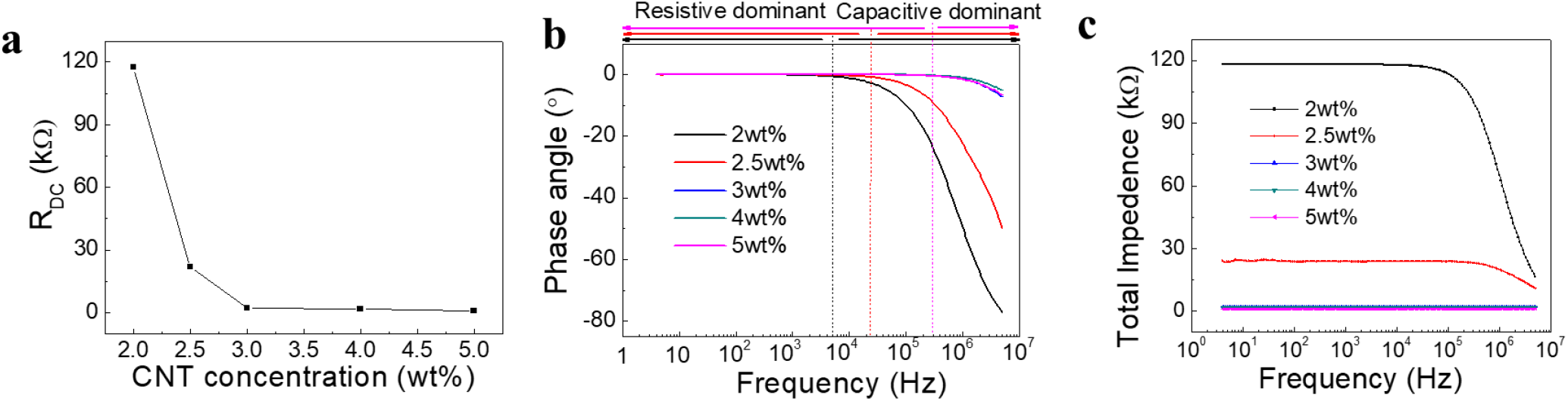
Electrical properties of the sponge-based sensor with different CNT contents. (a) Direct current resistance (R_DC_). Frequency-dependent (b) phase angle, and (c) total impedance of the CNT/PDMS sponges.

### Static pressure sensing performance of the device

According to the electrical property investigation above, the CNT/PDMS sponge-based sensor does not comprise solely resistive or capacitive materials. The sensors with 2wt% and 3wt% CNT content have the two typical different electrical properties. Therefore, the impedance response of the sponge-based sensor with 2wt% and 3wt% content to pressure were measured. In this study, different static pressures were applied using different weights and the measurement setup was shown in Figure 4**a**. The impedance parameters are dependent on the measurement frequency. To investigate the effects of measurement frequency on sensor sensitivity, impedance responses to different pressures were measured at different frequencies. The total impedance (Z) contains a real (resistance, Z_re_ = Z*cosθ) and imaginary component (reactance, Z_im_ = Z*sinθ). Figure 4**b**∼**d** present the total Z response and the Z_re_ and Z_im_ at 2wt% CNT and various pressures (27.9 Pa and 0.1, 0.5, 1.5, and 2.5 kPa) at 1, 2.5, and 5 MHz, respectively. The rate of change in Z_re_ is the highest but remains relatively low (<0.17). Figure 4**e**∼**g** show the sensitivities of the 2wt% CNT/PDMS sensor regarding of Z, Z_re_, and Z_im_. Sensitivity was calculated using the equation 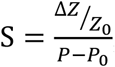, where ΔZ represents the change in impedance (or its real/imaginary part) in response to external pressure P, and Z_0_ is the initial impedance (or its real/imaginary part) of the pressure sensor. The curves can be delineated into two linear segments in the pressure ranges of 27.9 Pa–0.2 kPa and 0.2–2.5 kPa with all measurement frequencies. When CNT concentration is 2wt%, the response sensitivity of Z_re_ is relatively higher than that of Z and Z_im_. And it varied with different frequencies. The average sensitivities (i.e., the average of sensitivities at 1, 2.5, and 5 MHz) of Z_re_ were 0.51 kPa^−1^ and 0.0078 kPa^−1^ in pressure ranges of 27.9 Pa–0.2 kPa and 0.2–2.5 kPa, respectively, whereas the corresponding values for Z (Z_im_) were 0.28 kPa^−1^ (0.21 kPa^−1^) and 0.0045 kPa^−1^ (0.0047 kPa^−1^), respectively. As for 3wt% CNT concentration sponge, the corresponding data measured at 4 Hz, 1 MHz, and 5 MHz are shown in Figure 4**h**∼**j**, respectively. At 4 Hz, the sponges almost act as resistors, with no imaginary component. The change rate of Z_im_ is highest at 1 and 5 MHz, whereas that of Z and Z_re_ are similar at all measured frequencies because of the small phase angle even at 5 MHz. Unlike 2wt% CNT/PDMS sponge, the sensitivity of 3wt% CNT/PDMS is similar with different frequencies and different parameters (Z, Z_re_ and Z_im_), as shown in Figure 4**k**∼**m**. The overall average sensitivity (i.e., average values of all parameters at all frequencies) of 0.85 and 0.11 kPa^−1^ in the pressure ranges of 27.9 Pa–0.2 kPa and 0.2–2.5 kPa, respectively. The sensitivity data of the different test conditions were presented in Table **2**. It is obvious that 3wt% CNT content sensor has higher sensitivity than that of 2wt%. It can be concluded that the sensor performs better when it behaves more like a resistor (with the CNT content of 3wt% or above), and the sensing mechanism is piezoresistive. The sensitivity should be about 0.9kPa^-1^ (the value of Z_re_ sensitivity at 4Hz) when used in a DC circuit. Therefore, the sensor can be applied with a relatively simple measurement circuit (e.g., voltage divider circuit) and easily to be integrated with the MCU board which usually provide direct power supply.

**Figure 4.**
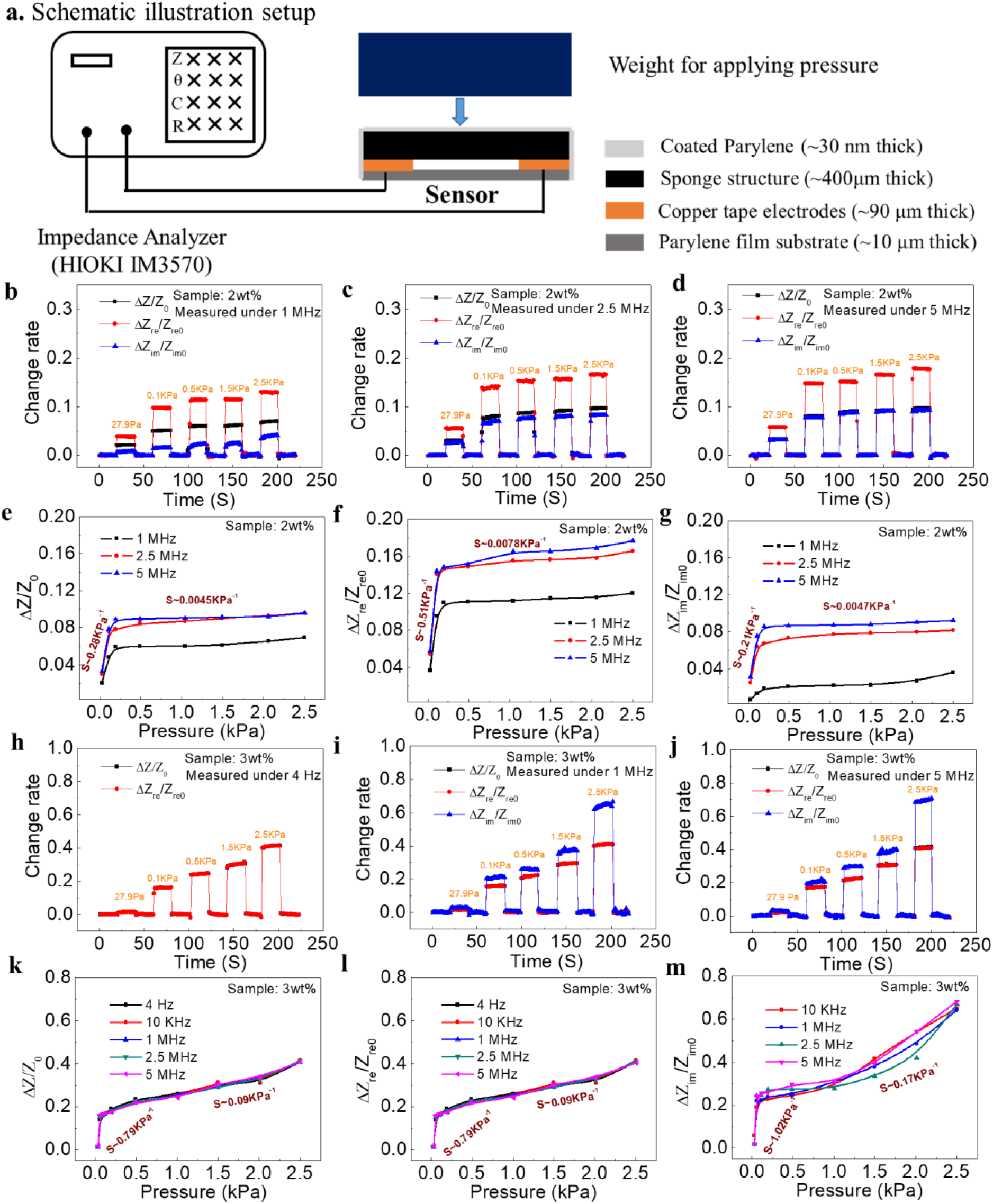
Impedance responses of CNT/PDMS sponge-based sensor with CNT concentrations of 2wt% and 3wt%. (**a**) Illustration schematic of measurement setup. (**b**∼**d**) Change rates and (**e**∼**g**) the corresponding sensitivity of Z, Z_re_, and Z_im_ at 1, 2.5, and 5 MHz for a sponge with 2wt% CNT concentration. (**h**∼**j**) Change rates and (**k**∼**m**) the corresponding sensitivity of Z, Z_re_, and Z_im_ at 4 Hz, 1 MHz, and 5 MHz for a sponge with 3wt% CNT concentration. (CNT: carbon nanotube; PDMS: polydimethylsiloxane; Z: impedance; Z_re_: real impedance component; Z_im_: imaginary impedance components.)

**Table 2.** Pressure sensitivity characteristics of total impedance (Z) and its real (Z_re_) and imaginary (Z_im_) components for a CNT/PDMS sponge with an approximate thickness of 400 µm and CNT concentration of 2wt% and 3wt%.

| Measurement<br>Frequency (Hz) | Segmented pressure sensitivity of Z, Z<sub>re</sub>, and Z<sub>im</sub> (obtained through linear fitting) |  |  |  |  |  |
| --- | --- | --- | --- | --- | --- | --- |
|  | 27.9 Pa–0.2 kPa |  |  | 0.2–2.5 kPa |  |  |
|  | Z | Z<sub>re</sub> | Z<sub>im</sub> | Z | Z<sub>re</sub> | Z<sub>im</sub> |
| 2wt% CNT/PDMS sponge-based sensor |  |  |  |  |  |  |
| 1MHz | 0.23 | 0.43 | 0.07 | 0.0042 | 0.004 | 0.0063 |
| 2.5MHz | 0.29 | 0.55 | 0.25 | 0.0069 | 0.0074 | 0.0053 |
| 5MHz | 0.33 | 0.54 | 0.32 | 0.0025 | 0.012 | 0.0025 |
| 3wt% CNT/PDMS sponge-based sensor |  |  |  |  |  |  |
| 4 Hz | 0.89 | 0.9 | -- | 0.08 | 0.08 | -- |
| 10 kHz | 0.78 | 0.78 | 0.83 | 0.09 | 0.09 | 0.19 |
| 1 MHz | 0.78 | 0.78 | 1 | 0.09 | 0.09 | 0.17 |
| 2.5 MHz | 0.8 | 0.8 | 1.16 | 0.09 | 0.09 | 0.15 |
| 5 MHz | 0.72 | 0.71 | 1.08 | 0.09 | 0.09 | 0.18 |

### Acoustic vibration frequency response of the device

The performance of the wearable sensor to high frequency vibration was also studied. With the above investigation, the sensor response was measured based on piezoresistive effect and a simple voltage divider circuit was applied (shown as the embedded graph in Figure 5**c**). The flexible sensor was attached to a vibration speaker (a commercial product of JBL Plus 3) and the speaker was controlled to vibrate at different frequencies by playing the corresponding audio. Meanwhile, a lase doppler vibrometer was applied to measure the actual vibration frequency and acceleration of the speaker (the detail testing setup is shown in Supplementary Figure S1). And the output voltage signal was recorded with the oscilloscope. Figure 5**a** presents the output voltage change when the speaker vibrates at different frequencies, and it shows the flexible sensor has a good response at high frequency vibrations and its amplitude is related with the vibration acceleration (abbreviated as ‘a’). The Fast Fourier Transform (FFT) analysis of the sensor output signal shows the flexible sensor can detect the fundamental vibration frequency correctly, and harmonics generated (Figure 5**b**). The sensitivity of the vibration sensor can be calculated by 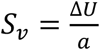 for piezoelectrical sensor[61]. As for our piezoresistive sensor, the sensitivity was calculated using 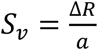. So, the sensor sponge-sensor has a sensitivity ranging from ∼2.4Ω/g to 4.4Ω/g, depending on the input frequency. The results show that its vibration sensitivity exhibits a slight increase and then become stable with the frequency increase (Figure 5**c**). Figure 5**d** shows the frequency components (FFT analysis) when the vibration speaker played with a series of piano scale A_3_, B_3_, C_4_, D_4_ and E_3_. Compared with the standard values, the flexible sensor shows a good accuracy and resolution when detecting these high-frequency vibrations. The sensor’s output signal was then converted to audio file and spectrum analysis taken, as shown in Figure 5**e**, fundamental frequency is detected, as well as the generated harmonics.

**Figure 5.**
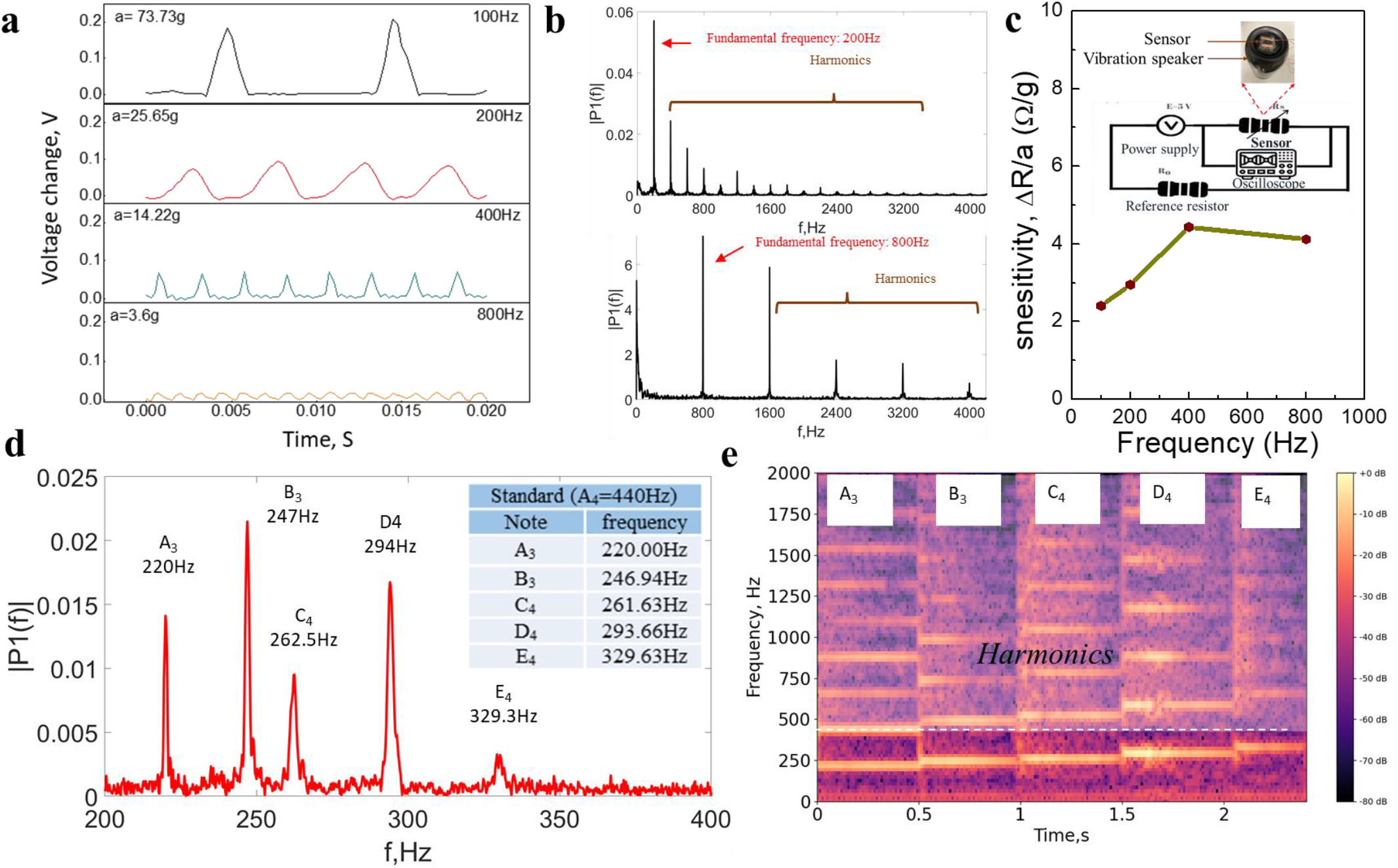
Vibration test of the fabricated flexible sensor using a vibration speaker. (**a**) Signal (voltage) change at different vibration frequency and acceleration. (**b**) The corresponding FFT analysis. (**c**) Vibration sensitivity with different frequency calculated based on the above test, and the measurement circuit embedded. (**d**) FFT analysis when the vibration speaker piano sound scale of A_3_, B_3_, C_4_, D_4_, E_4_ and which the corresponding spectrum graph.

### Detection of air directional flow and air vibration with the device

Above investigation and results show that the piezoresistive sponge-based sensor has a good sensitivity of both the static applied pressure and the high-frequency vibration under solid contact. Then, this sensitive flexible sensor was tested to detect the air pressure. Air movements including air directional flow and air vibration were studied. One is the air directional flow, where the air molecules flow in one direction without moving back and forth. The other is the air vibration, where the air molecules vibrate back and forth at some certain frequencies. The two pieces of electrodes of the sponge-based sensor was fixed to two separate pieces of glass with some distance to make the sensing area freestanding. A low cost, micro power consumption microphone preamplifier MAX4466 was used in this test, its circuit was shown in Figure 6**a**. MAX4466 is a mature product which is integrated with the electret microphone. Our developed sponge-based sensor can be easily integrated with this amplifying module by replacing the electret microphone and worked well. The air directional flow pressure was provided by an air gun blowing air to the sensor with constant velocity, and the air vibration was induced by the speaker playing a 315Hz sound in front of the sensor, Figure 6**b** shows the illustrations. The output voltage signal is varied with the resistance change of the sensor. Figure 6**c** shows the output electric signal of the air directional flow and air vibration, and Figure 6**d** shows the corresponding generated FFT (Fast Fourier Transform) plot. The results show that the air directional flow caused a irregular output signal during the record time and no definite frequency component with most energy was in the low frequency range. On the contrary, the air vibration caused a stable and regular output signal and the FFT analysis suggested that the sensor captured the 315Hz sound signal correctly. The frequency components at multiple of 315 Hz (e.g., 630 Hz, 945 Hz, 1260 Hz, etc.) were the generated harmonics since it is not a perfect sin wave (the 50 Hz signal was the noise caused by the power supply). The output electric signal was then converted to audio signal using a self-developed Python program since sound is actually the air vibration. And more information can be obtained using audio formant. Figure 6**e** presents the audio waveform of air directional flow and air vibration, and Figure 6**f** is the corresponding spectrogram. The amplitude of air directional flow audio was irregular while the air vibration produced the periodical audio amplitude. The audio spectrogram is the short-time Fourier transform of the input audio and represents the signal strength, or “loudness”, of a signal over time at various frequencies present the waveform which can also shows the energy levels vary over time. The spectrogram shows that the air directional flow mainly caused low frequency energy and vary with time. On the other hand, the energy of the air vibration caused signal focused on the 315 Hz and its harmonic frequencies (i.e., 630 Hz, 945 Hz, 1260 Hz…). In addition, the energy at these frequency keeps stable over time which is an important feature that differentiates it air directional flow signal. Another parameter power spectral density (PSD) which represents the spectral energy distribution per unit time shows that the energy of air directional flow decreases steadily with frequency increasing while the air vibration presented several peaks (315*n Hz, n=1,2,3…) over frequency (Figure 6**g**). Harmonic to Noise Ratio (HNR) measures the ratio between periodic and non-periodic components of the audio. Figure 6**h** shows the HNR value of the air directional flow and air vibration. The former (i.e., air directional flow) has a negative HNR value since it has almost no periodic component, while air vibration has a positive HNR value as it is the periodic signal. In summary, the air directional flow and air vibration caused signals are different in the aspects of both the time domine and the frequency domine.

**Figure 6.**
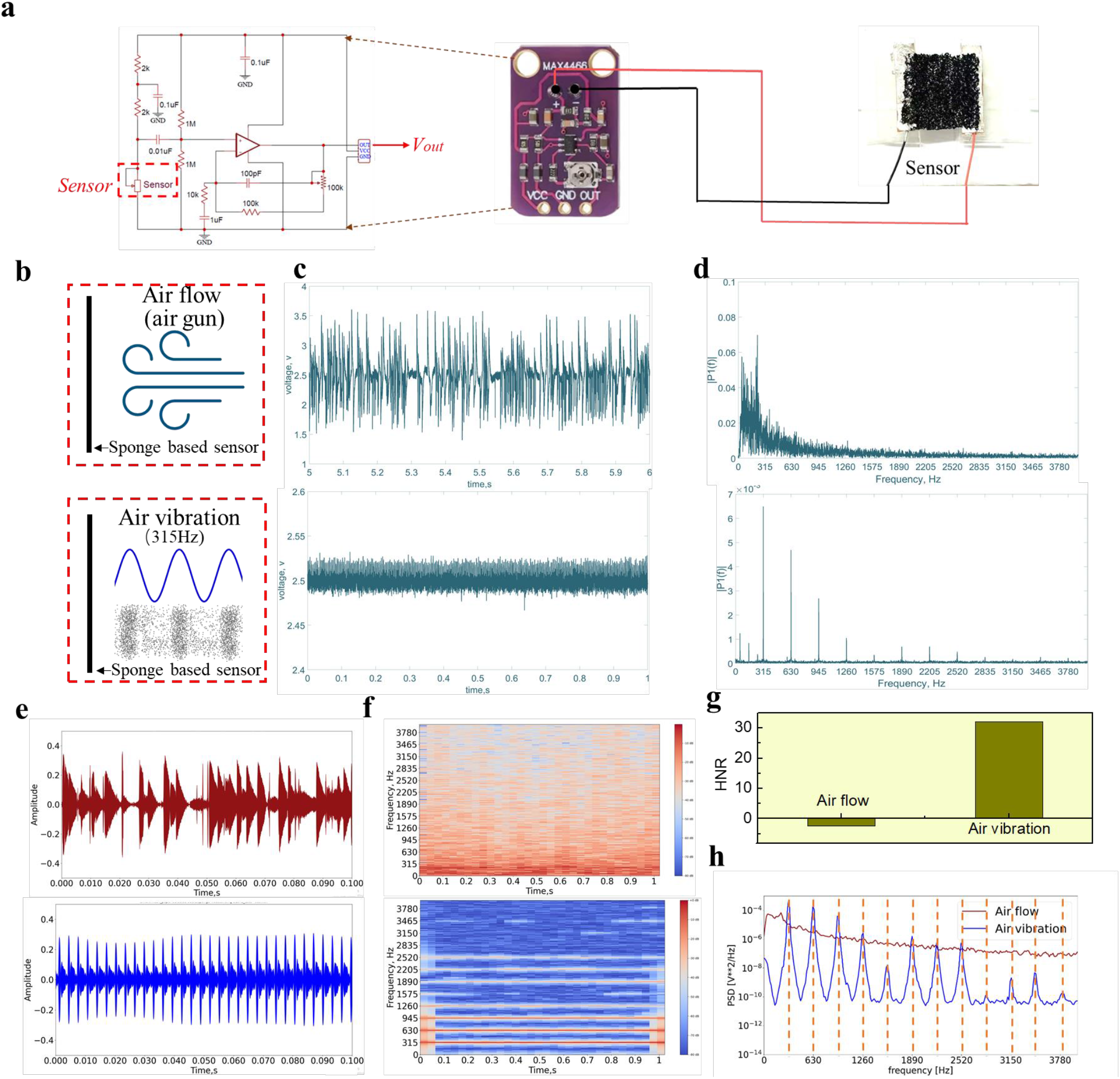
Detection of air directional flow and air vibration with the sponge-based sensor. (**a**) Measurement circuit. (**b**) Illustration of the setup of the sponge-based sensor detecting the air directional flow and air vibration signals. (**c**) Output electric signal caused by the air directional flow and air vibration. (**d**) The generated FFT plot. (**e**)The waveform of the audio converted by the air directional flow and air vibration signals. (**f**) Spectrograms of the air directional flow and air vibration audios. (**g**) PSD plot of the air directional flow and air vibration. (**h**) HNR value comparison of air directional flow and air vibration.

### Smart mask based on the flexible sponge-based sensor

The flexible thin sponge-based sensor has been demonstrated to have a high sensitivity regardless of static pressure and dynamic pressure. It can also detect the air movements including air directional flow and air vibration. So, an idea that integrating this sponge-based sensor to a commercial mask to measure human daily respiratory activities was proposed. As shown in Figure 7**a**, the sensor was fixed in front of the inside of the mask and keep freestanding to detect human breath, cough, and speech. Based on the production mechanism of breath [34], cough [38] and speech [39], the breath mainly involves the air directional flow process, cough involves both air directional flow and air vibration process, while speech mainly involves air vibration process along with some air directional flow. Therefore, these three respiratory activities should be detected by our sponge-based sensor integrated smart mask.

**Figure 7.**
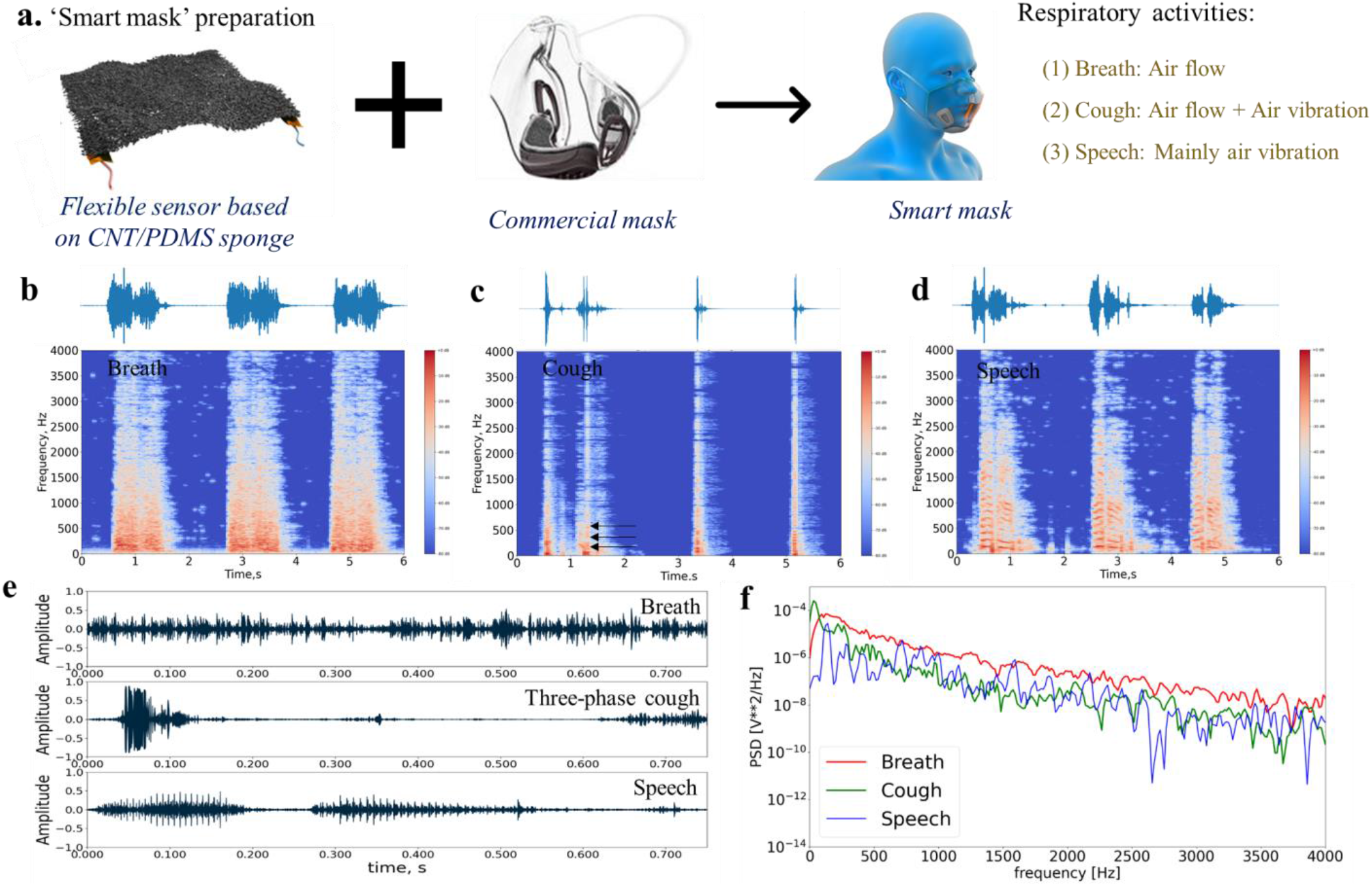
Smart mask to detect the human respiratory activities of breath, cough, and speech. (**a**) Illustration of the smart mask. (**b, c, d**) Waveforms and spectrograms of the audio signals of breath, cough, and speech. (**e, f**) Individual detailed waveform and PSD plot of the three kinds of respiratory activities.

Figure 7**b**∼**d** shows the waveforms and spectrograms of breath, cough and speech signals detected by the developed integrated smart mask. Figure 7**e** present the single clean signals of breath, three-phase cough and speech(‘robot’). It can be seen that the breath signal just like the air directional flow signal which has a irregular amplitude and mostly low frequency energy considering of a single clean breath signal. For continuous measurement signal (Figure 7**b**), the breath counting over time (breath rate) can also be obtained. The recorded cough signals consist of two-phase and three-phase cough, in which the third phase of the three-phase cough has the vibration features (short-time stability of frequency). Then the speech signal shows the obvious periodical vibration features. Its spectrogram shows the typical short-time stable, and it has the harmonic features. Similarly, coughs can be counted over time considering the continuous measurement (Figure 7**c**). Figure 7**f** gives the PSD curves of the three respiratory activities (i.e., breath, cough, and speech). The breath’s power spectra density decreases steadily with the frequency increasing, while for the speech signal, the value varies a lot and show significant peaks when decreasing along with the frequency. As for the cough, it performs between the breath and speech. The signal has a high energy in the very low frequency and then decreased as the frequency increasing with some obvious fluctuations. These results proved that the breath was detected as the air directional flow and speech was mainly detected as the air vibration while the cough had the features of both air directional flow and air vibration. In addition, the audio of the speech was heard to be right the ‘robot’, which indicates that our sensor can sense the subtle air vibrations. To make our developed smart mask portable and applicable, an ESP32 based wireless module was designed and prepared to sample the output signal and transmit the data to PC via WIFI protocol. The test results (Figure S2) show that the similar signals of breath, cough and speech can be obtained using this self-developed wireless device compared with the data recorded by the oscilloscope.

### Smart mask based on the flexible sponge-based sensor

To demonstrate our smart mask’s capability in monitoring people’s daily respiratory activities, 31 human subjects were recruited to perform breathing, coughing, and speaking while wearing the smart mask. The audio signals were acquired using an acquisition circuit based on the MAX4466 audio preamplifier module, and signals were sampled by both the oscilloscope and the self-developed wireless board. The following analysis was based on the data acquired from the two methods. Figure 8**a** shows the data process flow, in which the recorded voltage change signals were converted to audio files first and the segmentation work was performed manually based on the visualized waveform and spectrogram (as shown Supplementary Figure S3). Fifty-three features (see **Table1**), including HNR, PSD and etc, were extracted. The statistics for HNR value from the 31 subjects are shown in Figure 8**b**. The results suggest that speech has the highest and positive HNR value, which means most speech signals obtained were effective since they have the vibration features. The mean HNR value of breath is the lowest and negative because the breath signal has almost no periodical vibrations. As for the cough, its mean HNR value is between the speech and breath signals, which is reasonable because the first two phases of cough induced air flow while the third phase is voiced but not all coughs present this phase. The statistics for the data from thirty-one human subjects show consistency with the typical breath, cough, and speech signals in the main features, which suggests that our developed sponge-based smart mask has a good stability and robust performance. Principal Component Analysis (PCA) was used to simplify and compute the principal components of the extracted features, and Support Vector Machines (SVMs) classification algorithm was then applied to recognize the three different respiratory activities. Firstly, the classification was performed on the data of the individual 31 subjects while each of them has an average of total 108 pieces of data of the breath, cough and speech signals. The macro-recall (arithmetic mean of recall value for all the classes) and recall of each class (breath, cough, and speech) based on the classification results were calculated. Figure 8**c** shows the mean value of the recall of 31 human subjects, in which the mean recall value for each class is above 94% (i.e., 95.23%, 94.18%, and 96.29% for breath, cough and speech respectively). In addition, all the 31 subjects have a macro-recall above 90% (the maximum is as high as 100%) while the mean macro-recall value is about 95.23% (±3.36%). Then, the SVM classification was performed on the dataset from the total 31 human subjects (consist of 3368 pieces of data in total). The result has a macro-recall of about 91.43%, and the confusion matrix is shown in Figure 8**d**. In addition, Convolutional Neural Network (CNN) is also commonly used to analyze the respiratory sound with the input of spectrogram or Mel-spectrograms and have a good recognition accuracy of tasks of disease recognition [32, 33]. Therefore, further study on using CNN to perform the desired task was also carried. The whole process is shown in Figure 9**a**, MobileNet_v2 which consists of three convolutional layers, one pooling layer, seven bottleneck residual blocks and one fully connected layer was applied. Firstly, to obtain the same size of the input spectrograms, the segmented audio signals of different length was resampled and the spectrograms of the size of 256 (pixel) *256 (pixel) *3 (RGB channels) were generated. A random crop was then applied to modify the image shape to the MobileNet_v2 standard input size of 224*224*3. In the MobileNet_v2, convolutional layers apply a convolution operation to the input, then pass the calculation result to the next layer. The fully connected layer is the last layer, it would flat and compile the input to the shape of the final output. Rectified linear unit (ReLU) activation functions are used for the kernels in three convolutional layers and seven bottleneck residual blocks. Average pooling layers would perform the pooling operation that calculates the average value in the input. We utilized the cross-entropy loss function and the Adam optimization approach. We trained the network with a learning rate of 0.0001 with 200 epochs, the relationship between model accuracy with epoch is shown in Figure 9**b**. The last classification results based on the dataset of all thirty-one human subjects show a macro-recall of about 93.88%., as the confusion matrix shown in Figure 9**c**.

**Figure 8.**
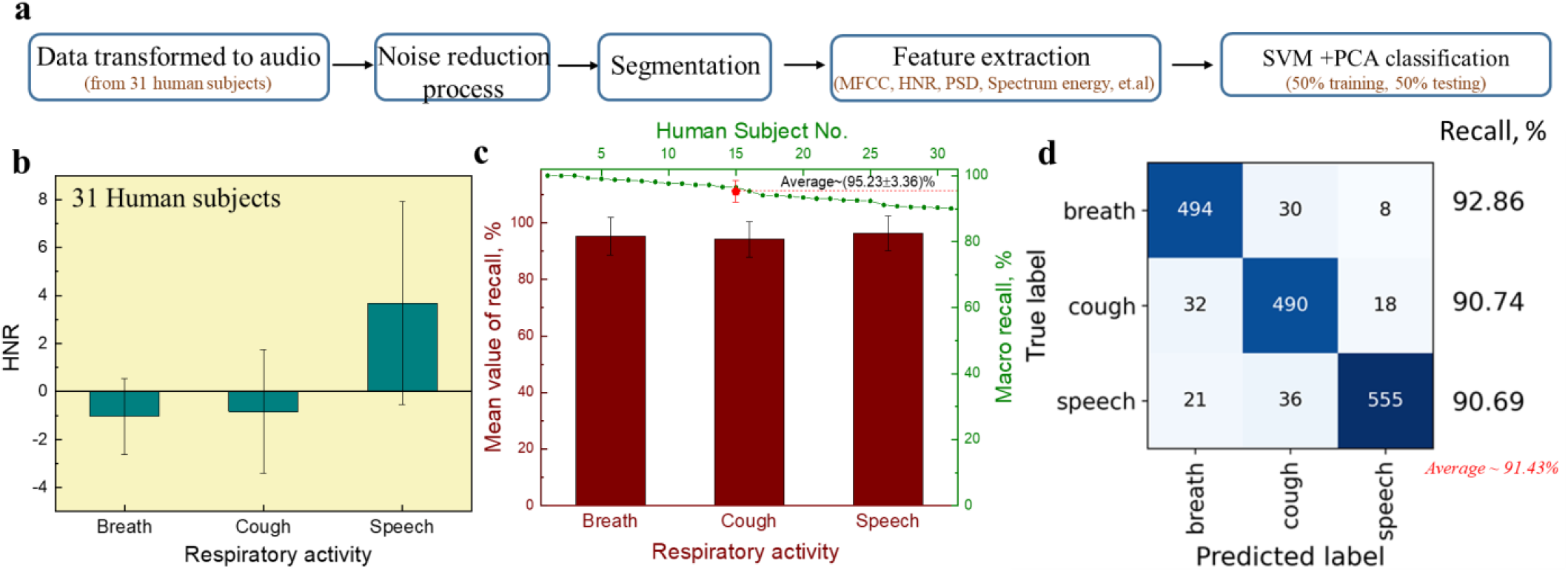
Data processing and classification. (**a**) Data processing flow. (**b**) The mean HNR value of breath, cough, and speech for thirty human subjects. (**c**) The mean value of the recall from the 31 individual subjects with PCA+SVM classification. (**d**) Confusion matrix of SVM classification results based on the dataset from all thirty-one human subjects.

**Figure 9.**
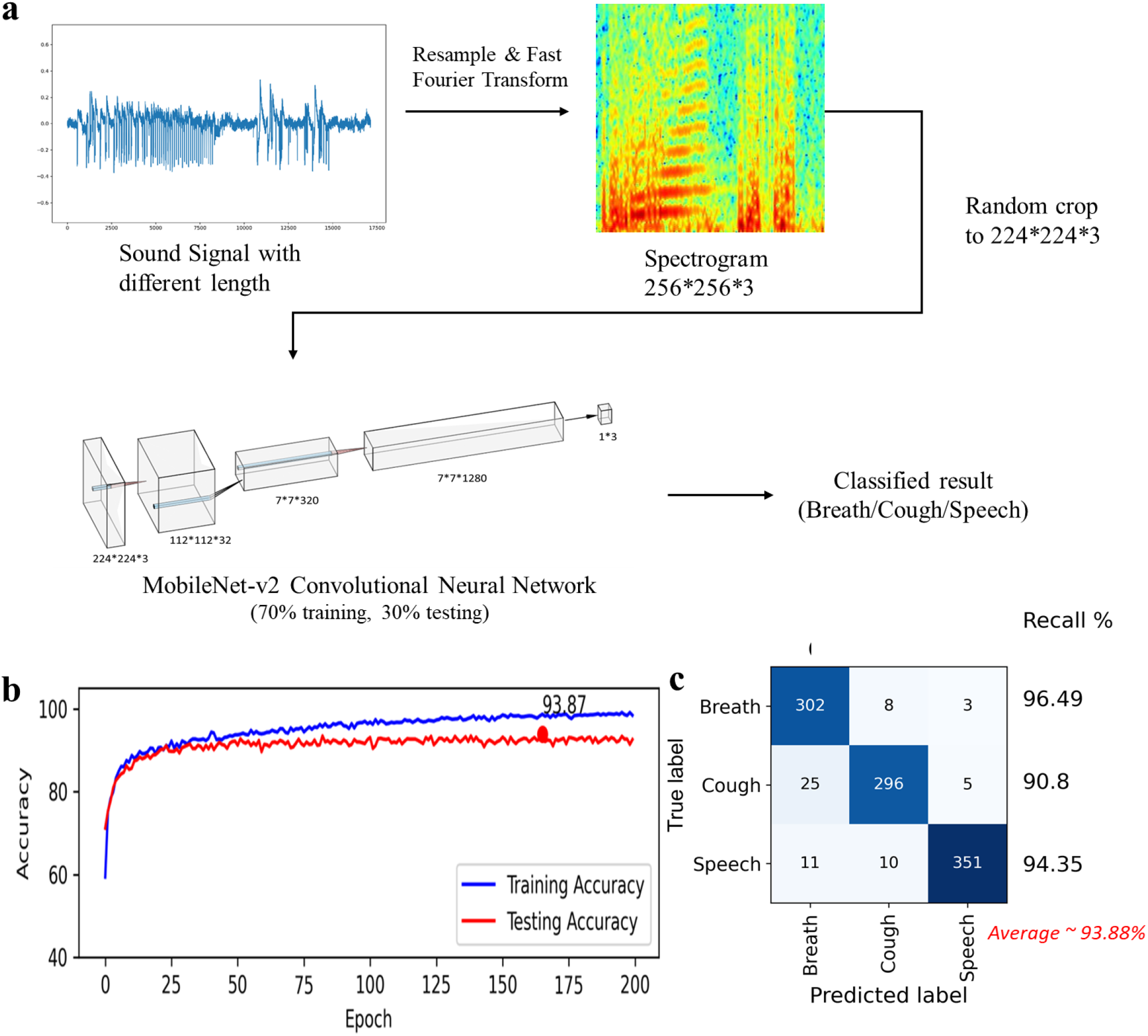
Classification using MobileNet_v2 CNN. (**a**) Process flow chart. (**b**) Relationship between model accuracy with epoch. (**c**) Confusion matrix of the classification results.

## Discussion and conclusion

Monitoring human daily respiratory activities using smart mask is a promising application. This research demonstrated a smart mask with integrating a nanomaterial-based high-frequency response acoustic pressure sensor to monitor human respiratory activities of breath, cough, and speech. The flexible pressure sensor was fabricated based on CNT/PDMS ultra-thin sponges with an excellent sensitivity in both static and dynamic pressure input, so this flexible pressure sensor has a wide application in monitoring of various human physiological signals whether at low-frequency or high-frequency. Therefore, the respiratory activities of breath, cough and speech can be all detected using the same ultra-lightweight sensor. To be applied inside the mask, the sensor should also be harmless to human body and work stably in high humidity environments. The sensing material (i.e., CNT/PDMS nanocomposites) has been proved to be biocompatible and no free CNTs observed during use [54]. And a layer of 30 nm thick parylene-c which is also biocompatible [62] and resistant to water vapor [63] was coated on the surface of the sensor to make it work well even in a humid environment and further prevent free CNTs from falling off. Figure S4 shows the contact angle measurement with liquid/volume of water/∼5μL on the sponge-based sensor. Therefore, this developed flexible sensor can be integrated inside the mask to detect the respiratory activities which is accompanied by the production of water vapor safely.

The respiratory activities mainly involve airflow and air vibration. In order to fulfill the monitoring respiratory activities at a certain distance from our mouth to nose, the developed sensor was verified of its ability to sense the different air movements (i.e., air directional flow and air vibration). For the smart mask preparation, the sensor should be kept freestanding between the mask’s inner surface and human face and fixed inside the mask for better detection which can also help effectively avoid the sound attenuation effect of the mask in high frequency region (i.e., above 1kHz). Experiments on human subject wearing this smart mask show that it can detect and differentiate three common respiratory activities including breath, cough, and speech. The breath signal was mainly due to the air directional flow, while cough signal included both air directional flow and air vibration (when the third voiced phase present). After converting these signals to audio sound, the audio corresponding to breath is a noisy sound since only air directional flow was induced without air vibration which cause meaningful sound. Cough sound can sometimes be heard in the third voiced phase because it would induce air vibration. It is suggested that not only the cough counting can be recorded but also the details of the cough signal can be obtained and analyzed using this smart mask. As for the speech, it was detected mainly due to the air vibration generated and so the corresponding audio can be clearly recognized. In addition to ‘robot’, different words (including both English and Chinese) were also recorded and can be recognized. The generated waveforms and spectrograms of the different words are shown in supplementary Figure S5. Although air-conducted human voice has a high frequency harmonics of thousands of hertz, the information up to 3400Hz should be enough for communication since this is the upper limit of the telephone bandwidth used today [64]. The spectrograms in Figure S5 shows that the sensor can detect the information of about up to 4kHz. Therefore, this smart mask has the great potential to help improving the speech intelligence which is degraded by the barrier of face mask. However, there are still some air flows which ‘impact’ on the sensor so the speech sounds to have some ‘popping sound’ that usually occurs when a speaker gets too close to a microphone. Applying a blowout hood may help reduce this popping noise and improve the audio quality. The results of the developed smart mask detecting human breath, cough and speech suggests that their distinguishable features can be identified, so that different activities can be recognized using our classification algorithm.

In conclusion, a flexible acoustic wave sensor was fabricated based on CNT/PDMS composite sponges with a modified imprint technique to make it as thin as 400μm. The sensor performs well with the piezoresistive mechanism according to the electrical properties and sensitivity investigation which shows 0.85 kPa^-1^ and 0.11kPa^-1^ in the pressure range of 27.9Pa∼0.2kPa and 0.2kPa∼2.5kPa, respectively. The flexible sensor was also tested to detect the dynamic pressure of the frequency of 100∼800Hz, which has a vibration sensitivity of about 3.48Ω/g. In addition, this flexible sensor has shown the ability to sense air movements including air directional flow and air vibration which have different characteristics (represented by features used in classification). That is, air directional flow caused relatively irregular signals and energy mainly focus on low frequency range, while air vibration caused periodical signal and energy focus on the vibration frequencies (fundamental frequency and the corresponding harmonics). These results show that the flexible sensor can be used to detect the human respiratory activities by integrating with a commercial polycarbonate mask. It was demonstrated that our developed ‘smart mask’ can detect and differentiate three common respiratory activities including breath, cough, and speech. The classification result of the three activities suggested an average macro-recall of about 95.23% (with individual dataset of 31 human subjects) and 93.88% (with a dataset containing all the 31 human subjects). We envision that the smart mask developed in this project could be enhanced by developing them into IoT devices based on encrypted Wi-Fi mesh technology, which will allow multiple nodes of masks to be distributed over a large social area and have the functions of self-organization and self-construction, and enable the masks to become multi-terminal, low power consumption, low cost, and high reliability IoT devices. We foresee other important applications in monitoring respiratory activities of subjects in high-risk or crowed environments, e.g., nursing homes, hospitals, and public transportation systems such as buses, trains and planes. Smart mask could also be further enhanced by adding other functionalities, i.e., by integrating it with off-the-shelf (e.g., see [14]) or custom-built sensors to monitor in real-time heart rate, blood oxygen saturation, blood pressure, and body temperature, etc., i.e., physiological parameters associated with symptoms of pneumonia caused by infectious viruses.

## Supporting information

Supporting Inormation

## Data Availability

All data produced in the present study are available upon reasonable request to the authors.

## Acknowledgement

This work was supported by the Shenzhen Municipality Science and Technology Innovation Commission (Grant No. SGDX2019081623121725) and Hong Kong Research Grants Council (Project No. 11204918 and 11216120).

## References

[1] A. Katzourakis, “COVID-19: endemic doesn’t mean harmless,” Nature, pp. 485–485, 2022.

[2] CDC. “COVID Data Tracker.” Atlanta, GA: US Department of Health and Human Services, CDC; 2022. https://covid.cdc.gov/covid-data-tracker/#variant-proportions.

[3] F. Wu et al., “A new coronavirus associated with human respiratory disease in China,” Nature, vol. 579, no. 7798, pp. 265–269, 2020.

[4] N. H. Leung et al., “Respiratory virus shedding in exhaled breath and efficacy of face masks,” Nature medicine, vol. 26, no. 5, pp. 676–680, 2020.

[5] K. K. Cheng, T. H. Lam, and C. C. Leung, “Wearing face masks in the community during the COVID-19 pandemic: altruism and solidarity,” The Lancet, 2020.

[6] Q. Wang and C. Yu, “The role of masks and respirator protection against SARS-CoV-2,” Infection Control & Hospital Epidemiology, vol. 41, no. 6, pp. 746–747, 2020.

[7] J. Howard et al., “An evidence review of face masks against COVID-19,” Proceedings of the National Academy of Sciences, vol. 118, no. 4, 2021.

[8] L. O. Gostin, I. G. Cohen, and J. P. Koplan, “Universal masking in the United States: the role of mandates, health education, and the CDC,” Jama, vol. 324, no. 9, pp. 837–838, 2020.

[9] K. L. Andrejko et al., “Effectiveness of face mask or respirator use in indoor public settings for prevention of SARS-CoV-2 infection—California, February–December 2021,” Morbidity and Mortality Weekly Report, vol. 71, no. 6, p. 212, 2022.

[10] The Associated Press. “Spanish PM calls for debate on treating COVID-19 as endemic.” 2022. Available at: https://abcnews.go.com/International/wireStory/spanish-pm-calls-debate-treating-covid-19-endemic-82175665.

[11] D. Thompsom, “California adopts nation’s 1st ‘endemic’ virus policyv.” The Associated Press, 2022. Available at: https://abcnews.go.com/Health/wireStory/california-adopts-nations-endemic-virus-policy-82965886.

[12] CDC. “Malaria’s Impact Worldwide.” Atlanta, GA: US Department of Health and Human Services, CDC; 2021. https://www.cdc.gov/malaria/malaria_worldwide/impact.html#:~:text=Nearly%20half%20the%20world's%20population,in%2087%20countries%20and%20territories.

[13] H. He et al., “Monitoring multi-respiratory indices via a smart nanofibrous mask filter based on a triboelectric nanogenerator,” Nano Energy, vol. 89, p. 106418, 2021.

[14] L. Pan et al., “Lab-on-mask for remote respiratory monitoring,” ACS Materials Letters, vol. 2, no. 9, pp. 1178–1181, 2020.

[15] R. Sabbadini, J. di Tocco, C. Massaroni, E. Schena, and M. Carassiti, “A smart face mask based on photoplethysmography for cardiorespiratory monitoring in occupational settings,” in 2021 IEEE International Symposium on Medical Measurements and Applications (MeMeA), 2021: IEEE, pp. 1–6.

[16] M. Wang et al., “Air-flow-driven triboelectric nanogenerators for self-powered real-time respiratory monitoring,” ACS nano, vol. 12, no. 6, pp. 6156–6162, 2018.

[17] Z. Zhongming, L. Linong, Y. Xiaona, Z. Wangqiang, and L. Wei, “Imperial graduate develops smart mask to track wearer’s respiratory patterns,” 2020.

[18] M. Lazaro, A. Lazaro, R. Villarino, and D. Girbau, “Smart Face Mask with an Integrated Heat Flux Sensor for Fast and Remote People’s Healthcare Monitoring,” Sensors, vol. 21, no. 22, p. 7472, 2021.

[19] W. Yang et al., “A breathable and screen-printed pressure sensor based on nanofiber membranes for electronic skins,” Advanced Materials Technologies, vol. 3, no. 2, p. 1700241, 2018.

[20] J. Zhong et al., “Smart Face Mask Based on an Ultrathin Pressure Sensor for Wireless Monitoring of Breath Conditions,” Advanced Materials, p. 2107758, 2021.

[21] N. Kim, J. L. J. Wei, J. Ying, H. Zhang, S. K. Moon, and J. Choi, “A customized smart medical mask for healthcare personnel,” in 2020 IEEE International Conference on Industrial Engineering and Engineering Management (IEEM), 2020: IEEE, pp. 581–585.

[22] M. Lazaro, A. Lazaro, R. Villarino, and D. Girbau, “Smart mask for temperature monitoring with LoRa backscattering communication,” in 2021 6th International Conference on Smart and Sustainable Technologies (SpliTech), 2021: IEEE, pp. 1–4.

[23] L. Ramazan, H. Karacali, A. Varol, U. Camli, and E. Yilmaz, “Fabrication and Characterization of Resistance Temperature Detector By Smart Mask Design,” 2021.

[24] M. H. Fakir and J. K. Kim, “Prediction of individual thermal sensation from exhaled breath temperature using a smart face mask,” Building and Environment, vol. 207, p. 108507, 2022.

[25] B. Varshini, H. Yogesh, S. D. Pasha, M. Suhail, V. Madhumitha, and A. Sasi, “IoT-Enabled smart doors for monitoring body temperature and face mask detection,” Global Transitions Proceedings, vol. 2, no. 2, pp. 246–254, 2021.

[26] N. V. R. Masna, R. R. Kalavakonda, R. Dizon, and S. Bhunia, “Smart and Connected Mask for Protection beyond the Pandemic,” in 2021 IEEE International Midwest Symposium on Circuits and Systems (MWSCAS), 2021: IEEE, pp. 676–679.

[27] N. V. R. Masna, R. R. Kalavakonda, R. Dizon, A. Bhuniaroy, S. Mandal, and S. Bhunia, “The Smart Mask: Active Closed-Loop Protection against Airborne Pathogens,” arXiv preprint 2008.10420, 2020.

[28] J. Korpáš, J. Sadloňová, and M. Vrabec, “Analysis of the cough sound: an overview,” Pulmonary pharmacology, vol. 9, no. 5-6, pp. 261–268, 1996.

[29] R. X. A. Pramono, S. A. Imtiaz, and E. Rodriguez-Villegas, “A cough-based algorithm for automatic diagnosis of pertussis,” PloS one, vol. 11, no. 9, p. e0162128, 2016.

[30] T. Nakajima et al., “Characteristics of patients with chronic cough who developed classic asthma during the course of cough variant asthma: a longitudinal study,” Respiration, vol. 72, no. 6, pp. 606–611, 2005.

[31] T. F. Quatieri, T. Talkar, and J. S. Palmer, “A framework for biomarkers of COVID-19 based on coordination of speech-production subsystems,” IEEE Open Journal of Engineering in Medicine and Biology, vol. 1, pp. 203–206, 2020.

[32] P. Bagad et al., “Cough against covid: Evidence of covid-19 signature in cough sounds,” arXiv preprint 2009.08790, 2020.

[33] K. K. Lella and A. Pja, “Automatic diagnosis of COVID-19 disease using deep convolutional neural network with multi-feature channel from respiratory sound data: cough, voice, and breath,” Alexandria Engineering Journal, vol. 61, no. 2, pp. 1319–1334, 2022.

[34] S. Cedar, “Every breath you take: the process of breathing explained,” Nursing Times, vol. 114, no. 1, pp. 47–50, 2018.

[35] T. Drugman et al., “Audio and contact microphones for cough detection,” arXiv preprint 2005.05313, 2020.

[36] C. Magni, E. Chellini, F. Lavorini, G. A. Fontana, and J. Widdicombe, “Voluntary and reflex cough: similarities and differences,” Pulmonary pharmacology & therapeutics, vol. 24, no. 3, pp. 308–311, 2011.

[37] J. Amoh and K. Odame, “Technologies for developing ambulatory cough monitoring devices,” Critical Reviews(tm) in Biomedical Engineering, vol. 41, no. 6, 2013.

[38] K. K. Lee et al., “Sound: a non-invasive measure of cough intensity,” BMJ open respiratory research, vol. 4, no. 1, p. e000178, 2017.

[39] Z. Zhang, “Mechanics of human voice production and control,” The journal of the acoustical society of america, vol. 140, no. 4, pp. 2614–2635, 2016.

[40] L. L. Mendel, J. A. Gardino, and S. R. Atcherson, “Speech understanding using surgical masks: a problem in health care?,” Journal of the American Academy of Audiology, vol. 19, no. 09, pp. 686–695, 2008.

[41] D. D. Nguyen et al., “Acoustic voice characteristics with and without wearing a facemask,” Scientific reports, vol. 11, no. 1, pp. 1–11, 2021.

[42] A. Goldin, B. Weinstein, and N. Shiman, “How do medical masks degrade speech perception,” Hearing review, vol. 27, no. 5, pp. 8–9, 2020.

[43] R. M. Corey, U. Jones, and A. C. Singer, “Acoustic effects of medical, cloth, and transparent face masks on speech signals,” The Journal of the Acoustical Society of America, vol. 148, no. 4, pp. 2371–2375, 2020.

[44] T. Rahne, L. Fröhlich, S. Plontke, and L. Wagner, “Influence of surgical and N95 face masks on speech perception and listening effort in noise,” Plos one, vol. 16, no. 7, p. e0253874, 2021.

[45] J. C. Toscano and C. M. Toscano, “Effects of face masks on speech recognition in multi-talker babble noise,” PloS one, vol. 16, no. 2, p. e0246842, 2021.

[46] T.-S. Dinh Le et al., “Ultrasensitive anti-interference voice recognition by bio-inspired skin-attachable self-cleaning acoustic sensors,” ACS nano, vol. 13, no. 11, pp. 13293–13303, 2019.

[47] J. J. Donai and R. M. Halbritter, “Gender identification using high-frequency speech energy: Effects of increasing the low-frequency limit,” Ear and Hearing, vol. 38, no. 1, pp. 65–73, 2017.

[48] H. A. Boyd-Pratt and J. J. Donai, “The perception and use of high-frequency speech energy: Clinical and research implications,” Perspectives of the ASHA Special Interest Groups, vol. 5, no. 5, pp. 1347–1355, 2020.

[49] E. N. MacDonald, M. K. Pichora-Fuller, and B. A. Schneider, “Effects on speech intelligibility of temporal jittering and spectral smearing of the high-frequency components of speech,” Hearing research, vol. 261, no. 1-2, pp. 63–66, 2010.

[50] Y. Maryn and N. Roy, “Sustained vowels and continuous speech in the auditory-perceptual evaluation of dysphonia severity,” Jornal da Sociedade Brasileira de Fonoaudiologia, vol. 24, no. 2, pp. 107–112, 2012.

[51] A. Tsanas, M. A. Little, P. E. McSharry, J. Spielman, and L. O. Ramig, “Novel speech signal processing algorithms for high-accuracy classification of Parkinson’s disease,” IEEE transactions on biomedical engineering, vol. 59, no. 5, pp. 1264–1271, 2012.

[52] R. J. Holmes, J. M. Oates, D. J. Phyland, and A. J. Hughes, “Voice characteristics in the progression of Parkinson’s disease,” International Journal of Language & Communication Disorders, vol. 35, no. 3, pp. 407–418, 2000.

[53] R. S. Irwin and J. M. Madison, “The diagnosis and treatment of cough,” New England Journal of Medicine, vol. 343, no. 23, pp. 1715–1721, 2000.

[54] J. H. Kim et al., “Simple and cost-effective method of highly conductive and elastic carbon nanotube/polydimethylsiloxane composite for wearable electronics,” Scientific reports, vol. 8, no. 1, pp. 1–11, 2018.

[55] J. N. Lee, C. Park, and G. M. Whitesides, “Solvent compatibility of poly (dimethylsiloxane)-based microfluidic devices,” Analytical chemistry, vol. 75, no. 23, pp. 6544–6554, 2003.

[56] W. D. Callister, Materials Science and Engineering: An Introduction; Solutions Manual to Accompany. Wiley, 1994.

[57] D.-Y. Jeon, H. Kim, M. W. Lee, S. J. Park, and G.-T. Kim, “Piezo-impedance response of carbon nanotube/polydimethylsiloxane nanocomposites,” APL Materials, vol. 7, no. 4, p. 041118, 2019.

[58] V. Panwar and K. Pal, “An optimal reduction technique for rGO/ABS composites having high-end dynamic properties based on Cole-Cole plot, degree of entanglement and C-factor,” Composites Part B: Engineering, vol. 114, pp. 46–57, 2017.

[59] L. Valentini, D. Puglia, E. Frulloni, I. Armentano, J. Kenny, and S. Santucci, “Dielectric behavior of epoxy matrix/single-walled carbon nanotube composites,” Composites Science and Technology, vol. 64, no. 1, pp. 23–33, 2004.

[60] A. Das, S. Sinha, A. Mukherjee, and A. Meikap, “Enhanced dielectric properties in polyvinyl alcohol–Multiwall carbon nanotube composites,” Materials Chemistry and Physics, vol. 167, pp. 286–294, 2015.

[61] Y.-F. Liu et al., “Spider-Inspired Ultrasensitive Flexible Vibration Sensor for Multifunctional Sensing,” ACS Applied Materials & Interfaces, vol. 12, no. 27, pp. 30871–30881, 2020.

[62] N. Stark, “Literature review: biological safety of parylene C,” Medical Plastic and Biomaterials, vol. 3, pp. 30–35, 1996.

[63] A. Qualtieri, F. Rizzi, G. Epifani, A. Ernits, M. Kruusmaa, and M. De Vittorio, “Parylene-coated bioinspired artificial hair cell for liquid flow sensing,” Microelectronic Engineering, vol. 98, pp. 516–519, 2012.

[64] C. Liu, Q.-J. Fu, and S. S. Narayanan, “Effect of bandwidth extension to telephone speech recognition in cochlear implant users,” The Journal of the Acoustical Society of America, vol. 125, no. 2, pp. EL77–EL83, 2009.

