## Supporting Inormation for "A Wide-bandwidth Nanocomposite-Sensor Integrated Smart Mask for Tracking Multi-phase Respiratory Activities for COVID-19 Endemic"

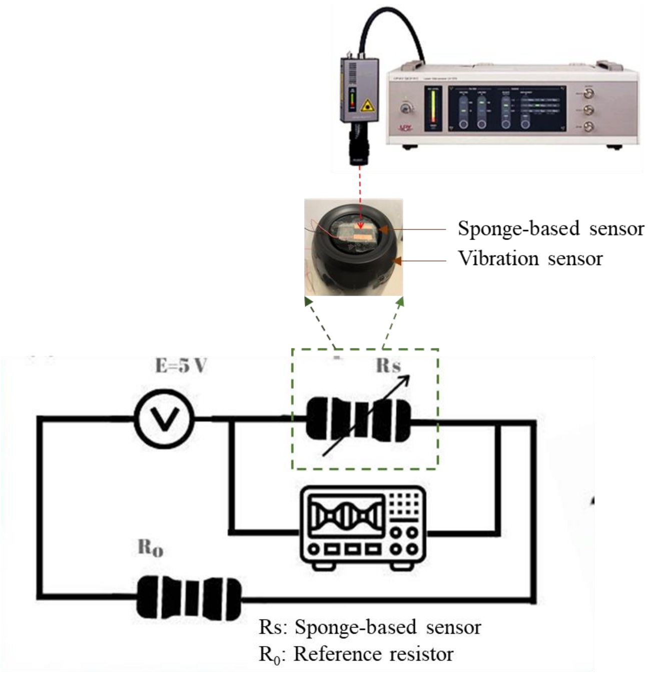


**Figure S1** Set up (including the measured circuit) of the sponge-based sensor detecting vibration of different frequency.

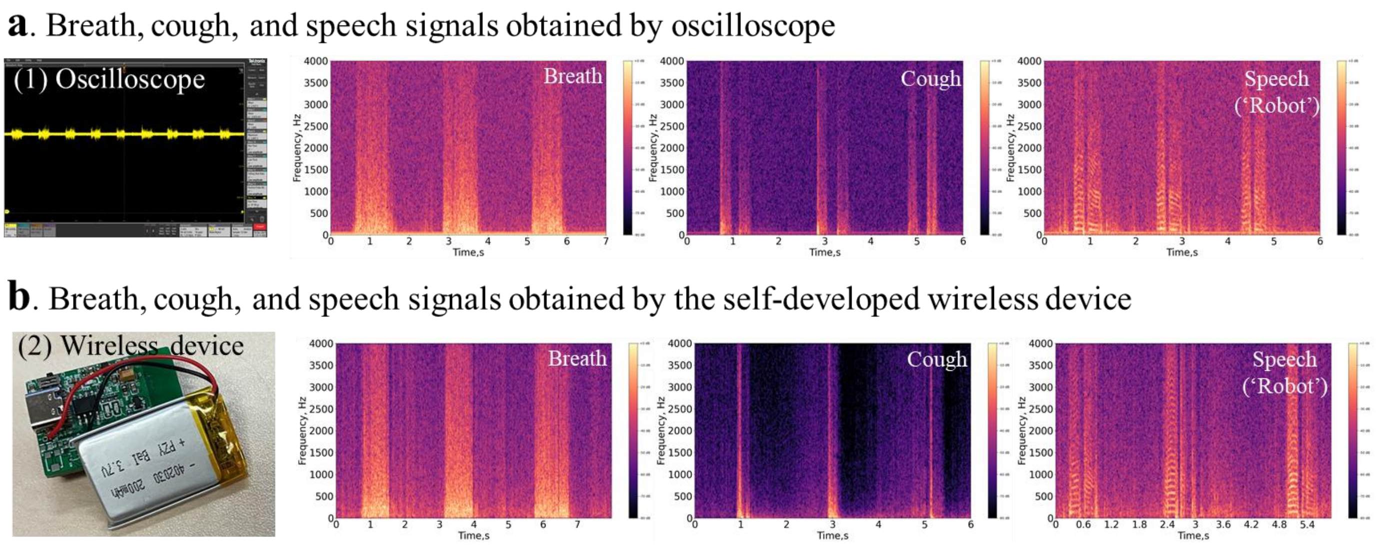


**Figure S2** Breath, cough, and speech ('robot') signals recorded with wearing the 'smart mask' with the data obtaining by: (a) oscilloscope and (b) the self-developed wireless device.

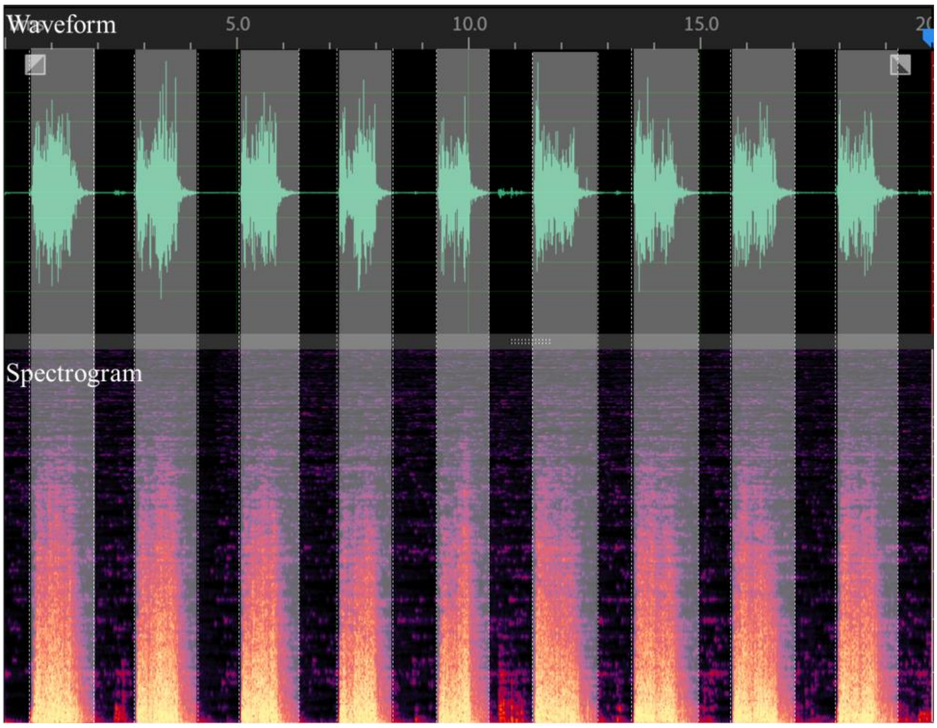


**Figure S3** Manual segmentation based on the visualized waveform and spectrogram

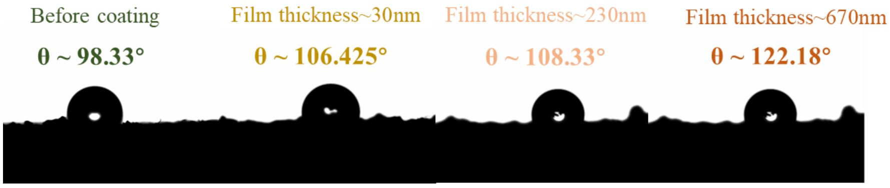


**Figure S4** Contact angle (θ) measurement (liquid/volume: water/~5μL)

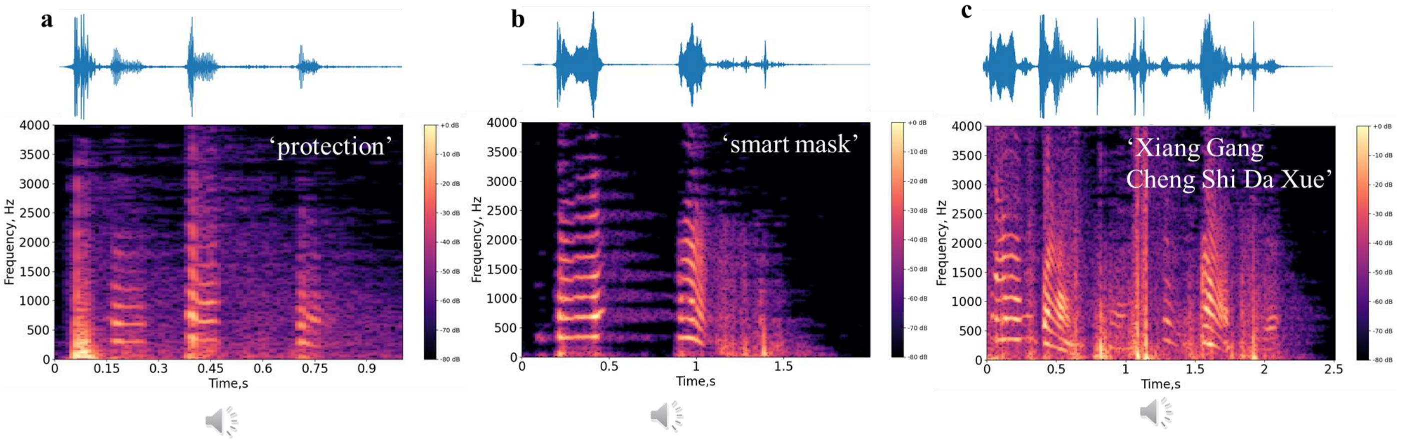


**Figure S5** Waveforms and spectrograms of the different words/phrases of (a) 'protection', (b) 'smart mask' and (c) Chinese 'Xiang Gang Cheng Shi Da Xue' detected by the 'smart mask'
